# Bayes Factors for Two-group Comparisons in Cox Regression with an Application for Reverse-Engineering Raw Data from Summary Statistics

**DOI:** 10.1101/2022.11.02.22281762

**Authors:** Maximilian Linde, Jorge N. Tendeiro, Don van Ravenzwaaij

**Author notes:** Correspondence concerning this article should be addressed to: Maximilian Linde, GESIS - Leibniz Institute for the Social Sciences, Cologne, Germany,.

## Abstract

The use of Cox proportional hazards regression to analyze time-to-event data is ubiquitous in biomedical research. Typically, the frequentist framework is used to draw conclusions about whether hazards are different between patients in an experimental and a control condition. We offer a procedure to compute Bayes factors for simple Cox models, both for the scenario where the full data are available and for the scenario where only summary statistics are available. The procedure is implemented in our “baymedr” R package. The usage of Bayes factors remedies some shortcomings of frequentist inference and has the potential to save scarce resources.

---

The biomedical literature is filled with studies in which two conditions are compared on some kind of outcome measure. A common example is a clinical trial in which the goal is to determine the efficacy of a therapeutic agent over a placebo or an already existing medication [e.g., 17, 29, 64]. The outcome measure can be continuous; an example would be symptom severity. Alternatively, the outcome measure can be dichotomous, as in studies that examine the mortality of patients following a medical procedure [see, e.g., 24, 57]. Sometimes, not only the sheer absence or presence of some event is relevant, but also the time until that event happens, which is often called the survival or failure time [37]. For instance, in order to judge the effectiveness of some form of oncological treatment, it is of interest to know how long terminally ill cancer patients survive after receiving the treatment, and at which time there is an increased or decreased risk of death.

Time-to-event data are typically analyzed using *survival analysis* [see 11, 12, 18, 19, 22, 37, 39, for excellent overviews]. Usually, researchers use the frequentist statistical framework for survival analyses. This, however, has several disadvantages: First, it is impossible to quantify evidence *in favor* of the null hypothesis [e.g., 60] of equal survival between conditions. The reason for that is that a non-significant finding can occur due to low power or a truly absent effect; the two possibilities cannot be disentangled [2, 47, 73]. Second, stopping data collection based on interim results (e.g, *p*-value already reached threshold or *p*-value did not yet reach threshold) is highly problematic because it increases the probability of having a false positive result [1, 59, 68].

We offer a procedure and easy-to-use implementation for hypothesis testing for survival analysis in the Bayesian framework. Specifically, we focus on *Cox proportional hazards regression* [henceforth called either Cox regression or Cox model; 23]. This allows directly contrasting the evidence for the null hypothesis ℋ_0_ that there is no effect with an alternative hypothesis ℋ_1_ that operationalizes that there is some effect; it also allows monitoring results and continuing or stopping data collection at will. Moreover, to the best of our knowledge, so far Cox regression can only be conducted when the full data are available. Oftentimes, however, it is relevant to reanalyze studies based on summary statistics reported in articles [e.g., in replications and meta analyses; e.g., 28, 67]. We propose a novel procedure that describes how data can be simulated based on summary statistics and how these simulated data can be used to subsequently conduct Bayesian hypothesis testing for Cox regression. We implemented both the process of data simulation and the process of Bayesian hypothesis testing in a software package [the “baymedr” R package; 52] that can be used by a wide audience of researchers, as we will illustrate.

The remainder of the article is organized as follows. First, we give an introduction to survival analysis in general and Cox regression in particular. Second, we explain how Bayes factors can be computed and interpreted and apply this to the special case of Cox regression. Third, we showcase how our “baymedr” software can be used to compute a Bayes factor when the full data are available. Fourth, we describe our procedure for the scenario where only summary statistics are available. Specifically, we describe how survival data can be simulated from summary statistics, we tune parameters for the data simulation process, and demonstrate how “baymedr” can be used to compute a distribution of Bayes factors for multiple simulated data sets. Fifth, we compare the performance (in terms of bias and variance) of our approach to an approximation approach advocated by [4].

## Survival Analysis

The time until the event of interest occurs is often called *survival time* or failure time. Typically, the survival time is only known for some participants (e.g., due to the study ending before the event is observed, participants withdrawing from the study, or failure to follow-up with participants), leading to right-censored observations [e.g., 37, 39, 48, 51]. One advantage of survival analysis is that it can handle this kind of incomplete data very well.

We denote the survival time as *T*. The probability that a participant is still alive after a particular time *t* is given by the *survival function*:

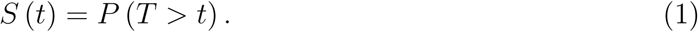

It is also informative to examine time periods of increased and decreased risk of failure. This is not immediately apparent in the survival function. The *hazard function* displays the instantaneous risk that the event happens in a narrow interval around a particular time *t* for participants who have survived until time *t* [cf. 18]:

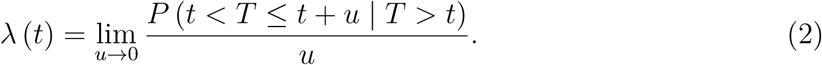

Various kinds of survival analyses exist for estimating *S* (*t*) and *λ* (*t*). The semi-parametric Cox regression [23] is used most commonly across all types of survival analyses. On the one hand, it is parametric because it assumes a multiplicative effect of the predictors on the hazard function (i.e., the assumption of proportional hazards); on the other hand, it is non-parametric because it does not impose any particular form on the hazard function. The focus of this paper lies exclusively on Cox regression.

### Cox Regression

In Cox regression, the data for each participant *i*, with *i* ∈ {1*, …, n*} where *n* represents the sample size, consists of the observed response *Y_i_* and an event indicator *δ_i_*, designating whether the event of interest occurred (1) or not (0).

We assume that we have one independent variable *x* that is dichotomous, indicating membership to one of two conditions (control = 0; experimental = 1). We refer to the combination of *Y*, *δ*, and *x* as the data *D* for a generic participant. The Cox model [23] is expressed as:

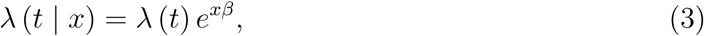

where *β* is the parameter we aim to estimate. Assuming that *x_i_* ∈ {0, 1}, the hazard ratio is:

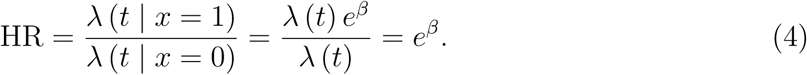

The estimation of *β* is based most often on maximum likelihood estimation. Commonly, either the original Cox partial likelihood [23], Breslow’s approximation to the true partial likelihood [13], or Efron’s approximation to the true partial likelihood [27] are used [e.g., 70]. We will exclusively use Efron’s method because it usually is most accurate, can handle tied survival times best, and is the default in several software packages; examples are the “survival” R package [69, 70] and the “rms” R package [38].

Once the model is estimated, a confidence interval for *β* or HR can be computed. Alternatively, null hypothesis significance testing (NHST) in the form of a Wald test (for instance) is conducted. Rejection of ℋ_0_ is warranted when the resulting *p*-value is smaller than a predefined significance level (when *p < α*); when *p* ≥ *α* nothing can be concluded.

## The Bayes Factor

The use of NHST in biomedical research is ubiquitous [16] even though it has been criticized repeatedly [e.g., 8, 9, 21, 26, 31, 33, 34, 35, 55, 72, 75, 76, 77]. Null hypothesis Bayesian testing (NHBT) is an alternative to NHST that has some practical advantages. For instance, in contrast to NHST, NHBT allows quantifying the relative evidence in favor of ℋ_0_ [60, 73, 75, 76]. That way, the evidence for (or against) ℋ_0_ and ℋ_1_ can be directly compared. This possibility is important because it enables researchers to investigate whether a therapeutic agent is *not* working. Moreover, in contrast to NHST, NHBT enables researchers to monitor the data during data collection and stop or continue data collection as needed [59, 61, 62, 63, 68]. This might have the implication that fewer resources are wasted because neither too many nor too few cases are sampled [15]. In addition, the results of NHBT are easy to interpret and are arguably more in line with researchers’ questions compared to NHST.

The most common vehicle of NHBT is the *Bayes factor* [41, 42, 43, 45], which quantifies the relative probabilities of the data under ℋ_0_ and ℋ_1_. For example, a Bayes factor of BF_10_ = 1 indicates a perfect balance between ℋ_0_ and ℋ_1_, given the choice of prior and model. In contrast, BF_10_ = 10 suggests that the data are 10 times more likely under ℋ_1_ compared to ℋ_0_, given the choice of prior and model. Lastly, BF_10_ = 0.1 indicates that the data are BF_01_ = 1*/*BF_10_ = 10 times more likely under ℋ_0_ compared to ℋ_1_, given the choice of prior and model. Even though the Bayes factor is a continuous measure of evidence, several schemes have been proposed to classify Bayes factors into categories that represent qualitatively different degrees of evidence. Table 1 shows Bayes factor thresholds suggested by [43]. Later, [50] offered alternative interpretations for the thresholds established by [43]. An alternative classification scheme was proposed by [45], with the following thresholds:

**Table 1.** Bayes factor evidence thresholds with interpretations proposed by [43] and adaptations thereof proposed by [50]. All interpretations are in favor of ℋ_1_; the same interpretations in favor of ℋ_0_ apply to the inverse of the Bayes factors.

| $\text{BF}_{10}$ | Jeffreys (1961)[43] | Lee & Wagenmakers (2013)[50] |
| --- | --- | --- |
| 1 |  | No evidence |
| 1 – 3 | Not worth more than a bare mention | Anecdotal evidence |
| 3 – 10 | Substantial evidence | Moderate evidence |
| 10 – 30 | Strong evidence | Strong evidence |
| 30 – 100 | Very strong evidence | Very strong evidence |
| > 100 | Decisive evidence | Extreme evidence |

1 − 3: not worth more than a bare mention; 3 − 20: positive; 20 − 150: strong; *>* 150: very strong. Importantly, all of these schemes should merely be considered as rules of thumb. They should be considered with caution and properly adapted to the problem at hand.

The Bayes factor follows from applying Bayes’ rule to both ℋ_1_ and ℋ_0_ using the available data *D*, while assuming that ℋ_1_ and ℋ_0_ are the two only models of interest:

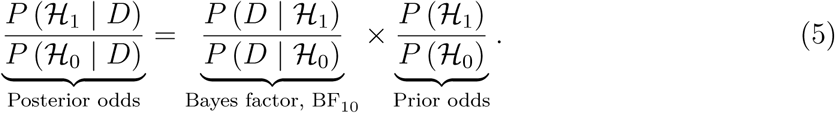

The prior odds reflect one’s initial beliefs about the probabilities of ℋ_1_ and ℋ_0_, the Bayes factor quantifies the relative probabilities of the data under ℋ_1_ and ℋ_0_, and the posterior odds reflect the relative probabilities of ℋ_1_ and ℋ_0_ after having observed the data. It can be seen in Equation 5 that the Bayes factor is independent of the prior odds. Therefore, when people hold different beliefs about the prior odds, they obtain different posterior odds but the Bayes factor remains the same.

The numerator of the Bayes factor in Equation 5 is computed by integrating the product of the prior distribution for the parameter of interest under ℋ_1_ and the likelihood function. In the case of Cox regression with one dichotomous independent variable, the parameter of interest is *β*. Consequently, the numerator of the Bayes factor in Equation 5 is:

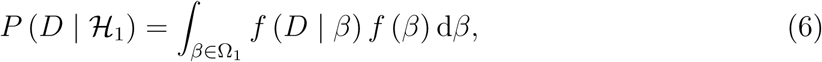

where Ω_1_ is the range of *β* parameter values under ℋ_1_. If a point ℋ_0_ is used, the denominator of the Bayes factor in Equation 5 is simply the density of the likelihood evaluated at the null value *β*_0_:

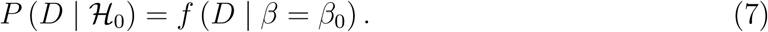

Even though the Bayes factor is independent of the prior odds, it is sensitive to the choice of prior for *β* [e.g., 30, 45, 66, 74].

The Bayes factor is computed the same way for either the full data or for data simulated based on summary statistics. The Bayes factor is:

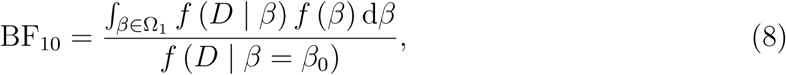

where *D* can refer either to the full data or the simulated data. Importantly, for our application of Cox regression, *f* (*D* | *β*) is equivalent to the natural exponent of Efron’s approximation to the true log partial likelihood.

## The Need for Methods for Full Data and Summary Statistics

The computation of Bayes factors can be challenging for applied researchers who do not have a firm background in Bayesian statistics and programming. Fortunately, multiple software packages are available that allow computing Bayes factors for various research designs. Examples are the R packages “BayesFactor” [56] and “baymedr” [52], and point-and-click software like “JASP” [40]. Moreover, for Bayesian parametric survival analysis, there is the “RoBSA” R package [3]. However, to the best of our knowledge, no software implementation currently exists that allows computing Bayes factors for Cox models. In addition to a module for computing Bayes factors with the full data set at hand, we also include a module that provides a highly accurate approximation of the Bayes factor when only summary statistics are available, for instance in the scenario where one reanalyses the results of a published study with only quantities published in the paper available.

In the next two sections, we showcase how researchers can use our “baymedr” R package [52] to compute Bayes factors for Cox models. In the first section, we focus on the situation where the full data are available. Subsequently, the second section focuses on the situation where only summary statistics are available. In that case, data must be simulated from the summary statistics; we explain how this is done, we tune data simulation parameters, and examine the bias and variance of the simulated Bayes factors.

The files with code for all computations can be found online (available at https://osf.io/37ut2/).

## Computing a Bayes Factor from the Full Data

We applied our approach for computing Bayes factors for Cox models to an empirical data set, as described in [5]. The goal of this double-blind, randomized, placebo-controlled trial was to determine the effectiveness of a therapeutic agent called Remdesivir for the treatment of the coronavirus disease 2019 (Covid-19). Participants were *n* = 1, 062 adults who were admitted to hospital due to Covid-19 infection. Participants were randomly assigned to a placebo condition (*n_c_* = 521) or a Remdesivir condition (*n_e_* = 541). The primary outcome was the time until recovery, which was conceptualized as patients either being dismissed from the hospital or remaining in hospital solely for the purpose of infection control.

[5] conducted a Cox regression to investigate the time until recovery. The authors used group membership (placebo vs. Remdesivir) as the independent variable and stratified by actual disease severity (severe disease vs. mild-moderate disease). In their supplementary material called “Protocol”, it is mentioned that a superiority ℋ_1_ is used (i.e., a one-sided ℋ_1_ with HR *>* 1) with a two-sided significance level of *α* = .05. More specifically, the authors hypothesized that patients receiving Remdesivir recover quicker than patients receiving a placebo. In contrast to our approach, the authors seem to have used Breslow’s instead of Efron’s approximation to the true partial likelihood. The authors conclude that “[p]atients in the [R]emdesivir group had a shorter time to recovery than patients in the placebo group” [5, p. 1816] and report a hazard ratio (i.e., recovery rate ratio) of HR = 1.29 together with a confidence interval of 95% CI = [1.12, 1.49] [see Table 2 in the article by 5].

Our reanalysis of [5] omitted the stratification by actual disease severity. Furthermore, we used Efron’s instead of Breslow’s approximation. The reason for these two deviations is that we have not implemented them in our “baymedr” R package. The resulting HR and the corresponding confidence interval are very close to [5]: HR = 1.312, 95% CI = [1.136, 1.514].

For the reanalysis we used our “baymedr” software, which can be downloaded and installed from GitHub using the “devtools” package [78] and loaded by typing the following into the R console:

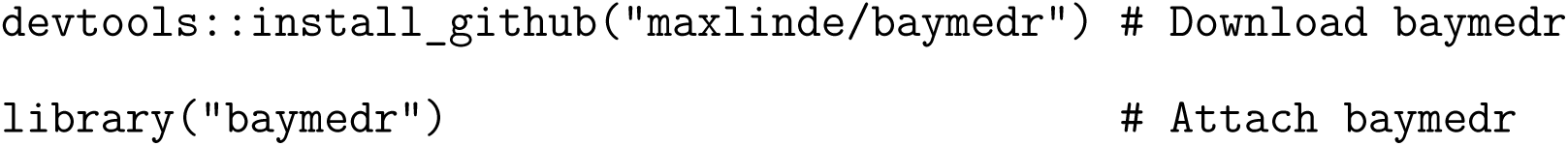

Using “baymedr”, we can compute a Bayes factor for the full data in [5]. The data must have the survival time, the event indicator, and the independent variable in that order as columns. We used a truncated (because we have a one-sided ℋ_1_) Normal prior for *β* with a mean of *µ* = 0 and a standard deviation of *σ* = 1:

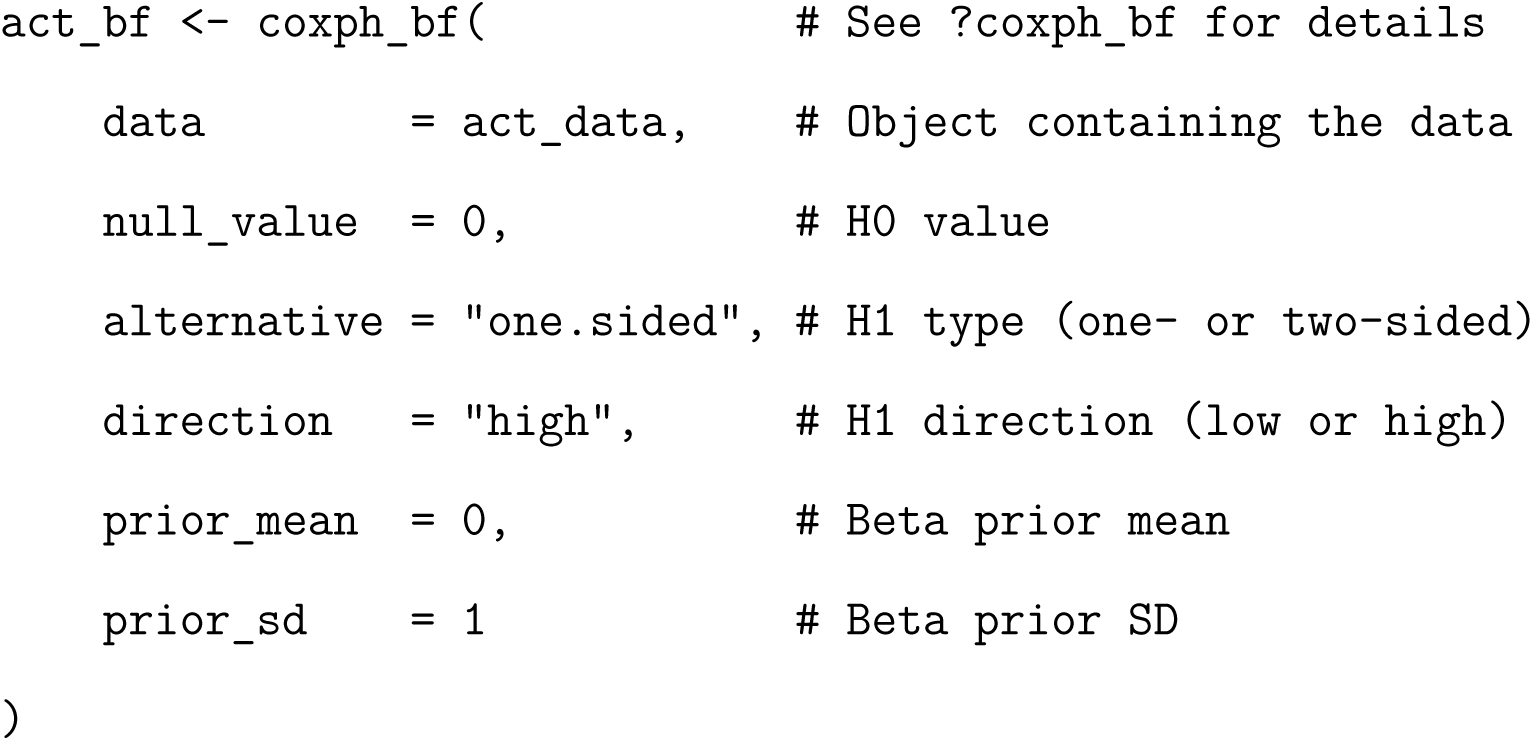

The obtained Bayes factor of BF_+0_ = 134.401 suggests that the data are 134 times more likely to have been generated under ℋ_1_ compared to ℋ_0_, given the choice of prior and model. Thus, according to approximate Bayes factor thresholds proposed by [43], we found decisive evidence (‘extreme’ according to [50] and ‘strong’ according to [45]) for the hypothesis that patients receiving Remdesivir recover quicker than patients receiving a placebo.

## Computing Bayes Factors from Summary Statistics

Sometimes, the full data might not be available. For example, when conducting a reanalysis of study findings, some researchers might not be allowed (e.g., for ethical reasons) or willing to share the full data. In these situations, it is paramount to be able to use summary statistics reported in the original manuscript to compute a Bayes factor.

When the full data are not available, an attractive alternative is to simulate data within the constraints of summary statistics that are known. Following such an approach, it is imperative to demonstrate that the simulated data are sufficiently constrained by the available summary statistics. To the best of our knowledge, our proposed approach for computing Bayes factors from summary statistics is unique and novel. Therefore, we deem our procedure an important contribution for verifying results of published studies. Our approach for data simulation is based on summary statistics that are commonly reported in scientific articles. Initially, we considered the following candidates:

- Sample sizes within each condition, *n_c_* and *n_e_*, respectively,
- Number of events within each condition, *v_c_* and *v_e_*, respectively,
- Maximum observed or maximum possible survival time, *t*_max_,
- Kaplan-Meier [KM; 44] median survival times within each condition, with corresponding confidence intervals, KM*_c_*, CI (KM*_c_*)*_LB_*, CI (KM*_c_*)*_UB_*, and KM*_e_*, CI (KM*_e_*)*_LB_*, CI (KM*_e_*)*_UB_*, respectively,
- Hazard ratio obtained from a Cox model [23], with corresponding confidence interval, HR, CI (HR)*_LB_*, and CI (HR)*_U B_*.

The first step for simulating data is to sample *n_c_* + *n_e_* responses *Y* drawn from a Uniform distribution:

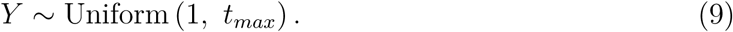

The choice of a Uniform distribution is arbitrary. Any other probability function would be equally suitable; even sampling *n_c_* + *n_e_* times the same value would suffice. These generated responses are paired with an event indicator *δ*:

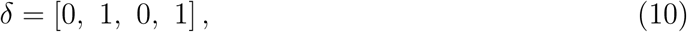

whose elements are repeated *n_c_* − *v_c_*, *v_c_*, *n_e_* − *v_e_*, and *v_e_* times, respectively. Lastly, the independent variable *x* is added:

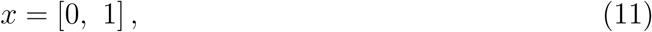

with the elements repeating *n_c_* and *n_e_* times, respectively. Then, *Y*, *δ*, and *x* form the preliminary simulated data *D_S_*, serving as a starting point for optimization.

Subsequently, summary statistics for the simulated data are computed. Possibilities are KM*_c_*, CI (KM*_c_*)*_LB_*, CI (KM*_c_*)*_UB_*, KM*_e_*, CI (KM*_e_*)*_LB_*, CI (KM*_e_*)*_UB_*, HR, CI (HR)*_LB_*, and CI (HR)*_U B_*. However, only a subset of the nine possibilities must be computed, namely those that are also reported in the article and that will therefore be used for data simulation.

The subsequent optimization procedure involves *n_c_* + *n_e_* parameters, which we collectively call *ξ*. Thus, each case *i* in *D_S_* is coupled with one parameter *ξ_i_* that must be estimated. At iteration *q*, *Y_i_* is computed as:

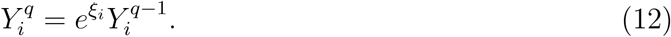

Here, *ξ_i_* is restricted to range between log 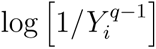 and log 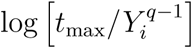; this ensures that the newly computed observed response 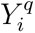 is not lower than 1 and not higher than *t*_max_. In essence, the optimization procedure attempts to adjust *Y* in a way such that interim summary statistics match the actual summary statistics. Let *E* be a vector of all known summary statistics and *O* be a vector (in fact, a function of *ξ* and *D_S_*) with the same kinds of summary statistics as *E* but computed from the simulated data *D_S_*. To estimate *ξ*, we iteratively minimize the following loss function:

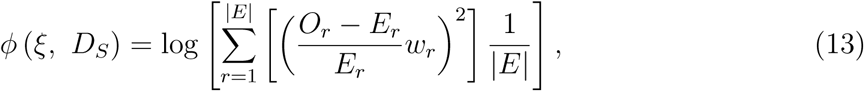

where |*E*| is the number of used summary statistics. *w* is a weight vector that we address below. In essence, we define the loss function as the log of the mean squared deviations between the observed and the expected summary statistics, scaled by the expected summary statistics (akin to the classical *χ*^2^ test statistic) and weighted by *w*. The scaling is done because the different kinds of summary statistics are on different scales and the weighting is done because different kinds of summary statistics might contribute more or less strongly to the accuracy of the resulting Bayes factors.

Remembering that *ϕ* (*ξ, D_S_*) is a (*n_c_* + *n_e_*)-variate function and that computing *O* involves complex formulas, it becomes clear that the loss function in Equation 13 is very difficult to differentiate. Therefore, gradient-based optimization techniques cannot be used; instead, we rely on a derivative-free optimization tool called *Particle Swarm Optimization* [PSO; 46, 65] to minimize Equation 13. A detailed treatment of PSO is beyond the scope of this article; we refer the interested reader to [20]. We implemented PSO in R [58] using the “psoptim()” function of the “pso” R package [7], keeping almost all default settings. Exceptions are the maximum number of PSO iterations and the allowed number of PSO iterations, which do not result in a decrease in the loss. The choice of our defaults is based on simulations, described below. Nevertheless, all arguments in the “psoptim()” function can be set as desired by the user.

### Tuning of PSO Parameters

For the tuning of some PSO parameters, we made use of three example data sets: the Kidney [54], Lung [53], and Colon [49] data sets that are available through the “survival” R package [69]. The Kidney data set provides times until infection after insertion of a catheter in kidney patients. The Lung data set describes survival times of patients with advanced lung cancer. Lastly, the Colon data set presents recurrence and death times in patients receiving adjuvant chemotherapy for colon cancer. Here, we chose to only examine death as an endpoint. For all three example data sets we used sex as the independent variable, with males being coded as 0 and females as 1. The Kidney, Lung, and Colon data sets have sample sizes of 76, 228, and 929, respectively. For the KM median survival times and HR we computed the corresponding 95% confidence intervals.

Importantly, we only used these data sets for the purpose of tuning the weights of summary statistics and the number of iterations in PSO. Therefore, no inferences from our results should be drawn.

#### Weights of Summary Statistics

The weight vector *w* in Equation 13 determines how influential certain summary statistics are in the computation of the loss function. We simulated 100 data sets for each of eight different sets of weights for the three example data sets. A Bayes factor was computed for each simulated data set. We used a Normal prior with a mean of *µ* = 0 and a standard deviation of *σ* = 1 for the *β* parameter. These simulated Bayes factors were then compared to the Bayes factor that we computed based on the full data set, using the same computational procedure as illustrated in the previous section.

The results are shown in Appendix A. For all three example data sets, a weight set in which the KM median survival times and the corresponding confidence intervals are not considered and HR is weighted twice as much as the corresponding HR confidence interval boundaries yields Bayes factors with the smallest variance. Moreover, there is almost no bias in the distribution of Bayes factors for the Lung and Colon data sets. A small amount of bias was found for the Kidney data set, which had the smallest sample size (see Figure A1). Using KM measures as well increases the variance and bias of the resulting Bayes factors. Consequently, it seems reasonable to ignore the KM estimates. In case the HR confidence interval is not mentioned in the original article, the results in Figures A1, A2, and A3 suggest that only using HR yields Bayes factors that are reasonable approximations to the true Bayes factor (Appendix B shows a follow-up analysis of the influence of specific weight combinations for HR, CI (HR)*_LB_*, and CI (HR)*_U B_* on the accuracy of the simulated Bayes factors, demonstrating that specific choices of weights do not matter). Due to these results, we henceforth only consider HR and the corresponding confidence interval as potential summary statistics. Further, using only HR and the corresponding confidence interval has the additional advantage that the maximum possible response time *t_max_* becomes irrelevant when simulating data.

#### Maximum Number of PSO Iterations

Another parameter of interest is the required number of PSO iterations for a satisfactory loss, bias, and variance of Bayes factors. This is especially important because the PSO algorithm is quite time-consuming. The higher the sample size, the larger the number of parameters in PSO and the longer the running time of PSO. To determine an approximate minimum number of PSO iterations, we simulated 100 data sets for each of six different maximum numbers of PSO iterations (i.e., 10, 30, 100, 300, 1, 000, and 3, 000) and computed the Bayes factor. This was repeated for each of the three example data sets. The PSO algorithm stops either when the maximum number of PSO iterations is reached or when no reduction in loss is obtained within one fifth of the maximum number of PSO iterations. Here as well, we used a Normal prior with a mean of *µ* = 0 and a standard deviation of *σ* = 1 for the *β* parameter.

The results are shown in Appendix C. Figure C1 indicates that a small number of PSO iterations is required when only using HR because the variance of the Bayes factors does not improve markedly when more than approximately 30 or 100 iterations are used. In contrast, Figure C2 suggests that when using both HR and the corresponding confidence interval the variance of the Bayes factors decreases the more PSO iterations are used. To obtain a reasonable trade-off between the running time of PSO and the Bayes factor variance, we recommend running between 100 and 300 iterations.

### Reanalysis of [5]

To demonstrate our proposed procedure for simulating data from summary statistics and computing one Bayes factor for each simulated data set, we again reanalyzed the study reported in [5]. But this time we will only used the summary statistics reported in their article. As in the previous section, our reanalysis of [5] omitted the stratification by actual disease severity and we used Efron’s instead of Breslow’s method. Due to these small deviations, our results (i.e., HR = 1.312, 95% CI = [1.136, 1.514]) are slightly different from the results reported in [5] (i.e., HR = 1.29, 95% CI = [1.12, 1.49]). We used our computed summary statistics as if they were the only results provided in [5].

Using the “baymedr” R package, we simulated 100 data sets based on the summary statistics as follows:

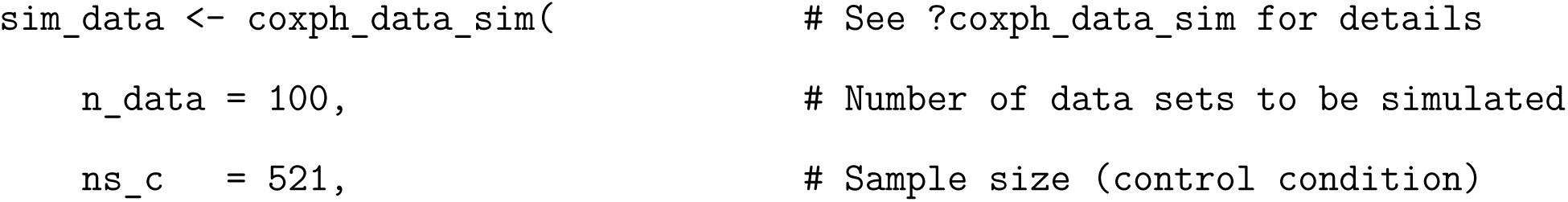
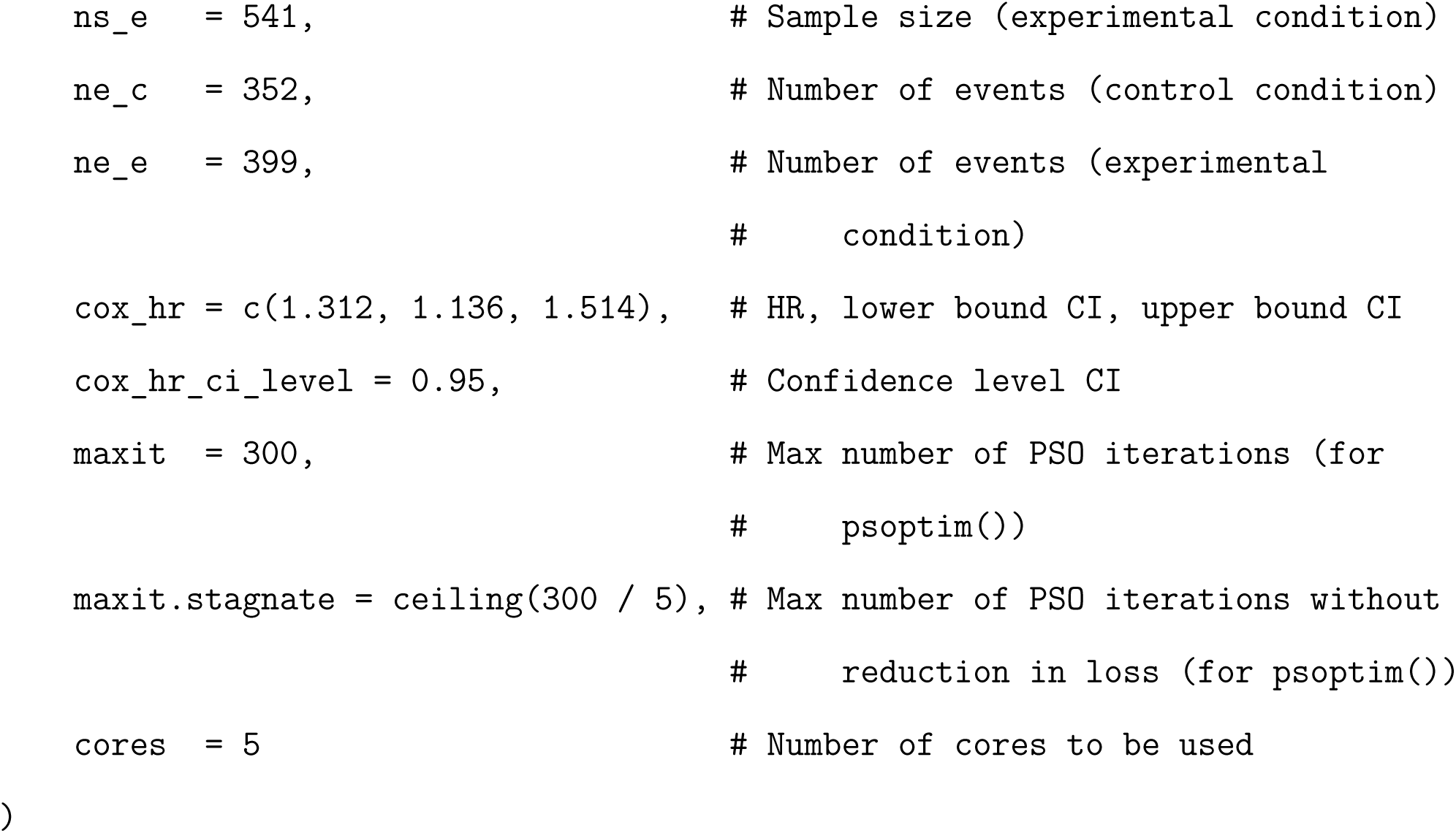

Subsequently, we computed one Bayes factor for each of the 100 simulated data sets. We used a truncated (because we have a one-sided ℋ_1_) Normal prior for *β* with a mean of *µ* = 0 and a standard deviation of *σ* = 1:

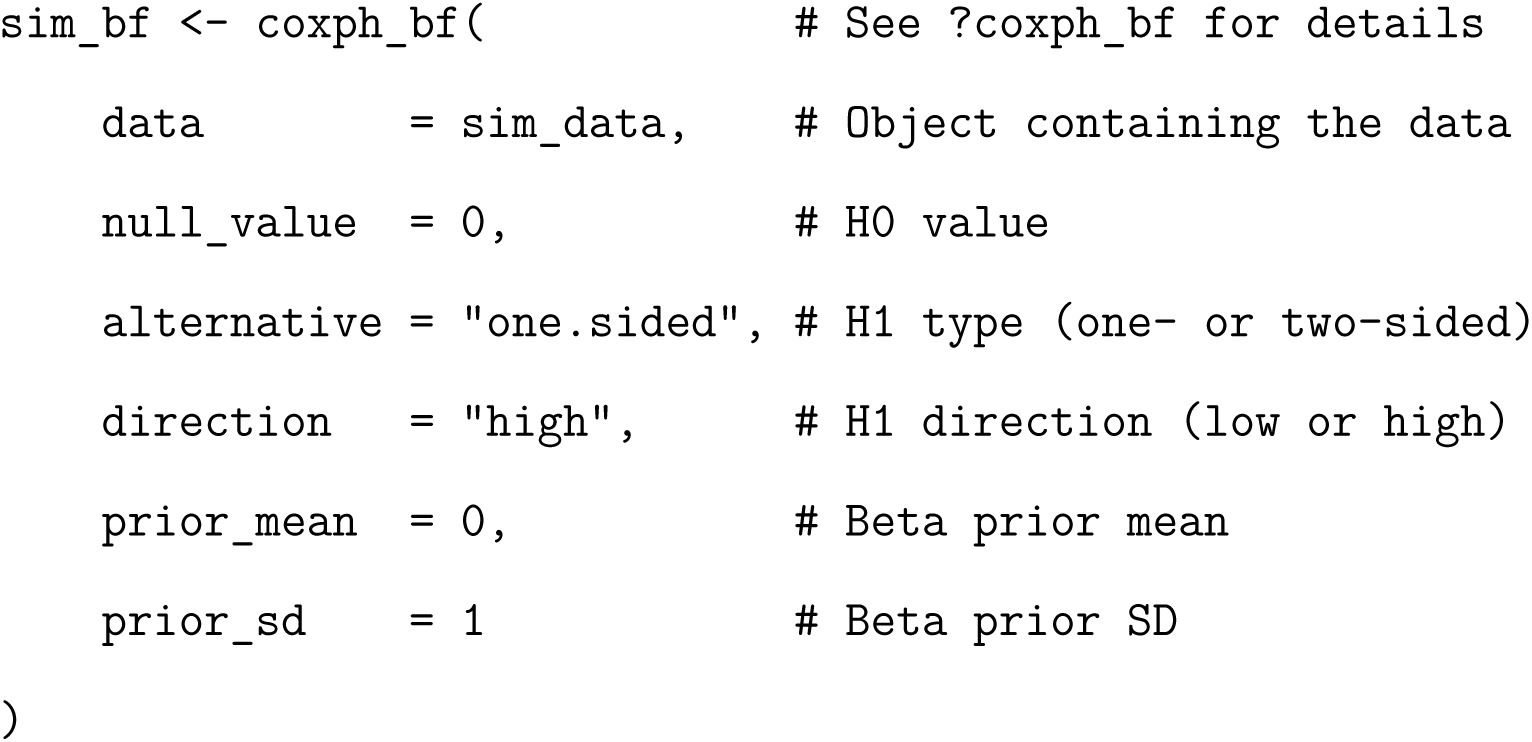

A histogram of the resulting Bayes factors can be found in Figure 1. The simulated Bayes factors range between BF_+0_ = 133.6 and BF_+0_ = 135.7, all providing ‘decisive’[43], ‘extreme’[50], or ‘strong’[45] evidence supporting the conclusion of [5] that Remdesivir seems to have a beneficial effect on the recovery of patients with Covid-19. The red vertical line represents the actual Bayes factor that was obtained in the previous section where we used the full data of [5]. As such, we can conclude that our approximate Bayes factor based on summary statistics is virtually unbiased (i.e., the red line is in the middle of the black histogram) and has very low variability (i.e., the histogram occupies a very limited range on the x-axis).

**Figure 1.**
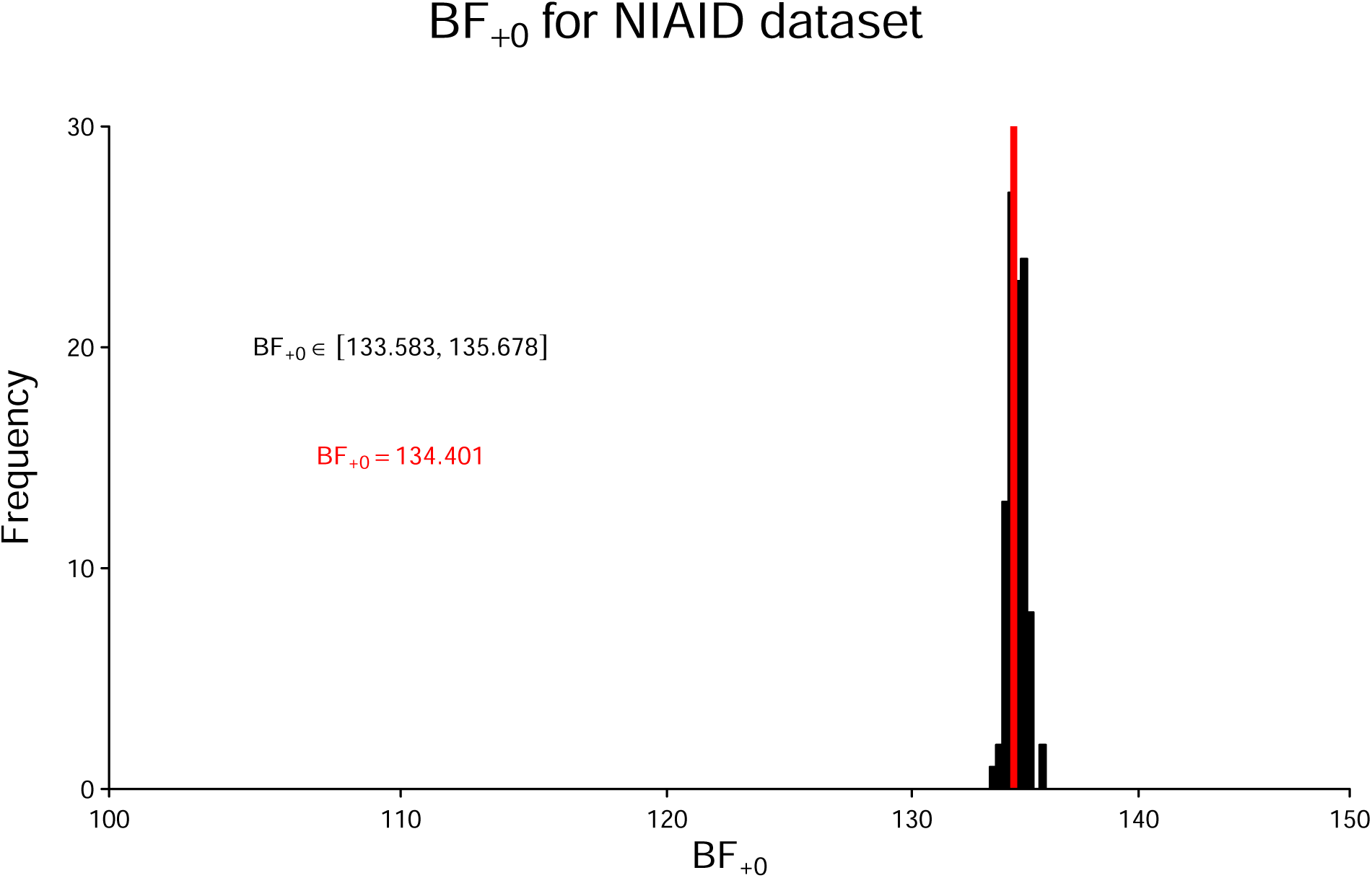
Distribution of BF_+0_ applied to 100 simulated data sets for the data set described in [5]. HR and its 95%CI are used for data simulation. The red vertical line represents BF_+0_ for the full data. See text for details.

## Comparison with Savage-Dickey Normal Approximation

In the following, we compare our procedure for computing Bayes factors with an alternative procedure that tries to approximate Bayes factors. [4] introduced a generic method that uses a Normal approximation of the likelihood function to compute a Bayes factor for various statistical designs. If ℋ_0_ is a point hypothesis, the Bayes factor is the ratio of the ordinate of the prior (i.e., density or height of the prior) and the ordinate of the posterior for the parameter of interest *β*, evaluated at the null value *β*_0_. This ratio is called the Savage-Dickey density ratio [e.g., 25]:

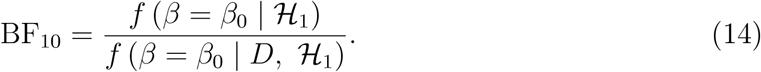

For this to work, only the maximum likelihood estimate and the corresponding standard error of the underlying likelihood function of the respective statistical analysis must be known (*β̂* and *SE* (*β̂*), respectively). If the prior for the parameter of interest is defined as a Normal distribution with mean *µ* and variance *σ*^2^, a closed-form solution for the Bayes factor is available [cf. the last equation on p. 3 of 4]:

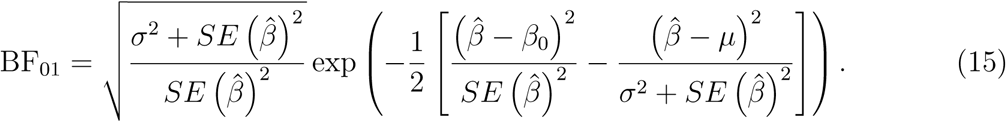

[4] use the examples of a two-sample *t*-test, a parametric survival analysis, and a meta-regression to demonstrate that their approximate Bayes factors are accurate and can be applied to a wide range of statistical models.

We investigated whether the method by [4] also yields accurate Bayes factors for semi-parametric Cox models and how they compare to the Bayes factors resulting from our method. This is important because the method by [4] provides a closed-form solution for computing Bayes factors from *β̂* and *SE* (*β*) directly. In other words, there is no need for simulating data from summary statistics, which makes their method time-efficient. As such, if both methods were to provide equally accurate Bayes factors, the method by [4] would be preferable. We refer the reader to Appendix D for a demonstration of typical run times of our method. We took the three example data sets mentioned before and computed one Bayes factor through our method and one Bayes factor through the method by [4], to mimic the situation where full data sets are available. Moreover, we simulated 100 data sets for each example data set using the corresponding summary statistics of the three data sets and computed one Bayes factor for each simulated data set using our method.

The results are shown in Figure 2, where the red vertical line represents the Bayes factor for the full data set computed using our method, the blue vertical line represents the Bayes factor resulting from the Savage-Dickey Normal approximation method advocated in [4], and the histogram shows Bayes factors from our method when using summary statistics. The Bayes factors resulting from the method by [4] are qualitatively similar to the true Bayes factors and can be used when a rough approximation is acceptable. However, when precise estimates are desirable, our method is preferable, both in the scenario where the full data set is available and in the scenario where only summary statistics are available. For the user it is a trade-off between accuracy and computation time. As shown in Figure 2, our Bayes factors are much more accurate (i.e., the distance between histograms and red lines is less than the distance between the blue lines and red lines). However, in the case where summary statistics must be used, our approach has a computation time that is orders of magnitude higher than the approach by [4] (see Appendix D).

**Figure 2.**
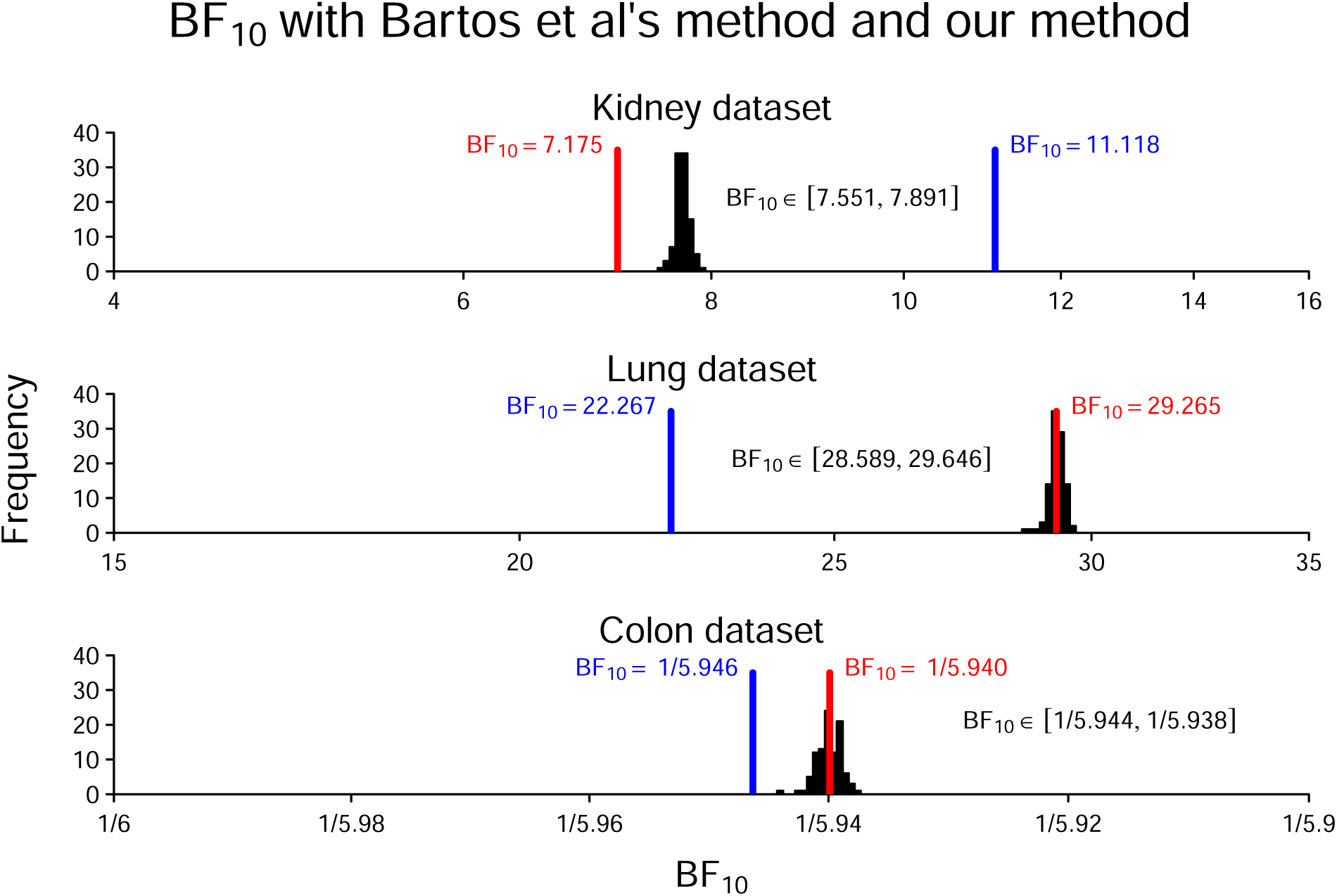
Distribution of BF_10_ applied to 100 simulated data sets for the Kidney, Lung, and Colon data sets using our approach. HR and its 95%CI are used for data simulation. The red vertical line represents BF_10_ for the full data set using our approach. The blue vertical line represents BF_10_ for the approximation by [4].

## Discussion

The analysis of time-to-event data is commonly applied in biomedical research and provides important insights into the effectiveness of therapies. Most often, Cox regression [23] is used to analyze these kinds of data and NHST is then applied in order to make inferences. As an alternative to NHST, we presented a procedure to compute Bayes factors for simple Cox models and offered the R package “baymedr” [52] as an easy-to-use implementation. “baymedr” can be used to compute a Bayes factor for full data and to simulate multiple Bayes factors based on summary statistics as reported in articles.

Our procedure for computing Bayes factors for Cox models is oriented towards analysis strategies that seem prevalent in the biomedical literature: the semi-parametric Cox regression comparing a treatment to a control (or different treatment) condition. The use of Bayes factors specifically allows the important contrast between evidence that an effect is present and evidence than an effect is absent and allows for optional stopping during data collection.

At the same time, many features and functionalities are still missing. For example, it would be desirable to also make the Cox partial likelihood [23] and Breslow’s approximation to the true partial likelihood [13] available. Moreover, the procedure implemented in “baymedr” should allow researchers to compute Bayes factors for more complex Cox models. This includes, for instance, allowing for more than one independent variable, whether it be discrete or continuous, and allowing for stratification. Such an extension to more than one independent variable is not straightforward, as it will not be possible to compute the Bayes factor through Gaussian quadrature. Instead, one of many more time-consuming approaches would have to be employed. For instance, the posterior distribution could be estimated through MCMC sampling [e.g., 10, 14, 32, 71]; the posterior samples could then be used to estimate the marginal likelihood through bridge sampling [e.g., 36].

## Conclusion

Cox proportional hazards regression is commonly used to analyze time-to-event data in biomedical research. Typically, the frequentist framework is used to make inferences. We provided a procedure for computing Bayes factors for simple Cox models that can be applied both to the full data set and to summary statistics. The latter could be considered especially important because it allows reanalyzing multiple existing studies to make judgments and decisions about the effectiveness of therapies. We offered “baymedr” [52], an R package that is aimed at all researchers desiring to compute a Bayes factor for their Cox regression.

## Data Availability

All data produced are available online at https://osf.io/37ut2/.

## Acknowledgments

We thank Merle-Marie Pittelkow for consulting us on which summary statistics are reported most often in articles that use a Cox proportional hazards regression.

## Appendix A

### Results for Weights of Summary Statistics

Figures A1, A2, and A1 show the bias and variance of the simulated Bayes factors based on different combinations of weights of summary statistics in the computation of the loss function for the Kidney [54], Lung [53], and Colon [49] data sets, respectively, from the “survival” R package [69]. See main text for details.

**Figure A1.**
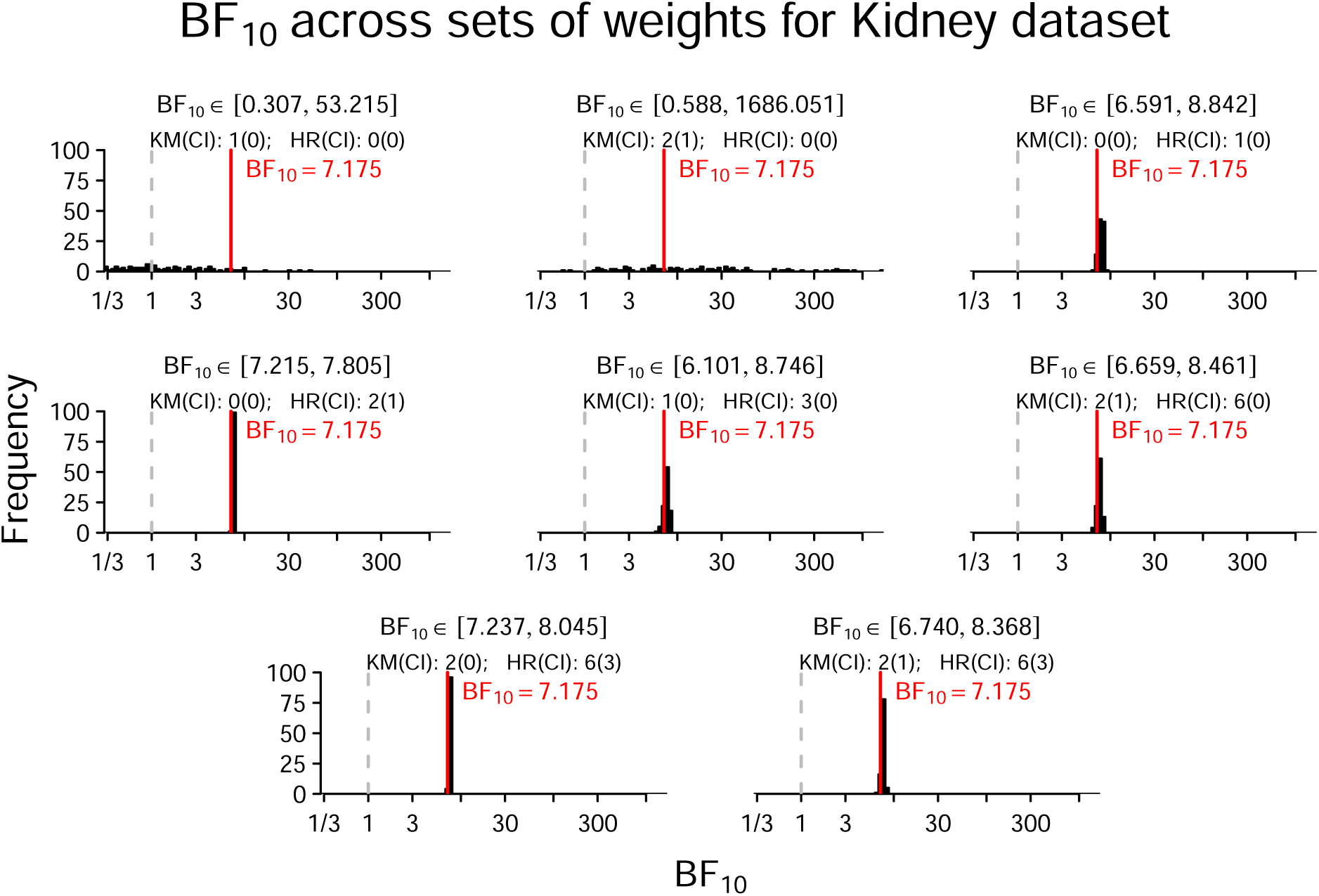
Distribution of BF_10_ for the Kidney data set. Panels display BF_10_ for 100 simulated data sets using different sets of weights for summary statistics. The specific weights are printed in each panel, where KM represents KM_c_ and KM_e_, CI represents CI (KM_c_)_LB_, CI (KM_c_)_UB_, CI (KM_e_)_LB_, and CI (KM_e_)_UB_, HR represents HR, and CI represents CI (HR)_LB_ and CI (HR)_UB_. The red vertical line represents BF_10_ for the full Kidney data.

**Figure A2.**
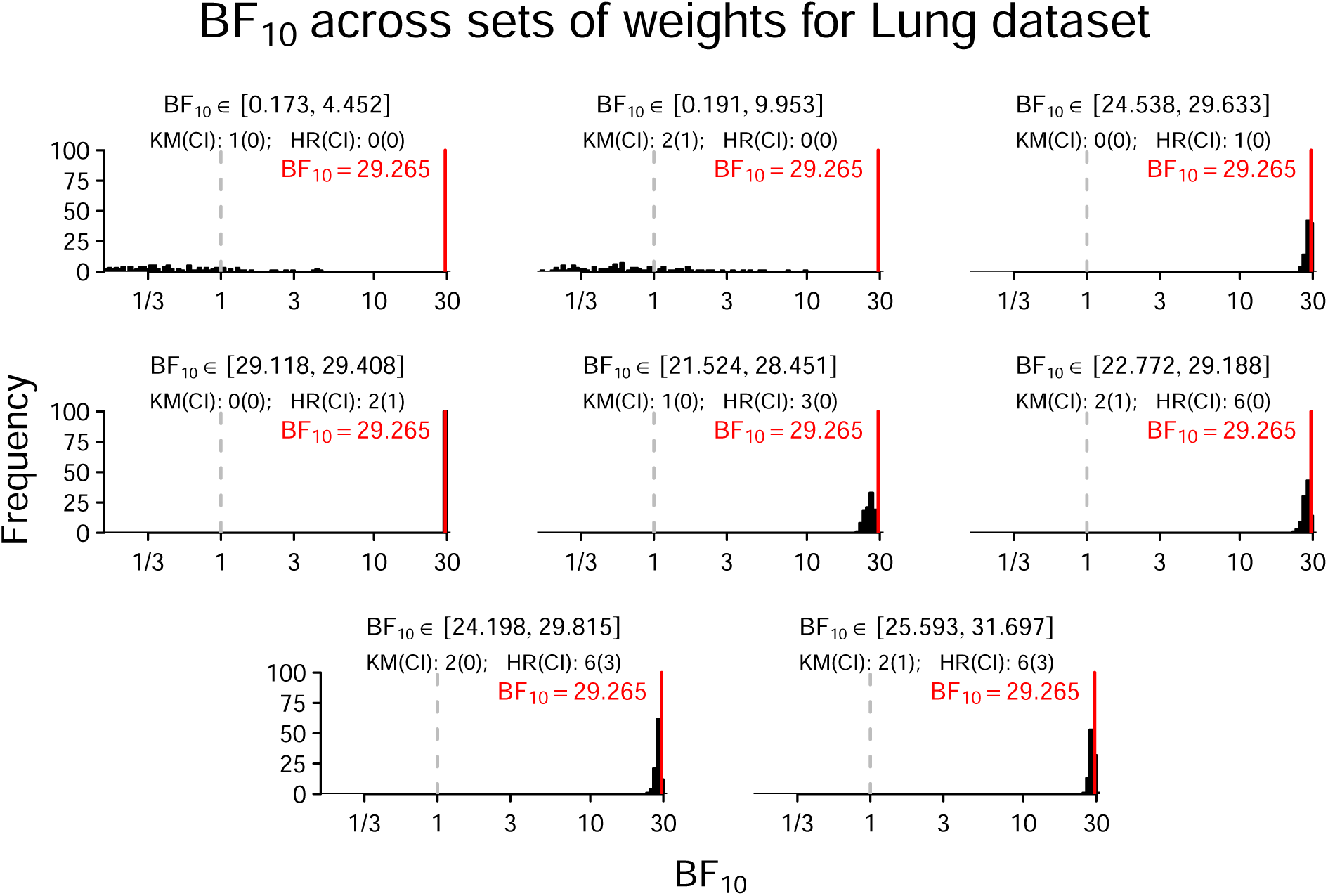
Distribution of BF_10_ for the Lung data set. Panels display BF_10_ for 100 simulated data sets using different sets of weights for summary statistics. The specific weights are printed in each panel, where KM represents KM_c_ and KM_e_, CI represents CI (KM_c_)_LB_, CI (KM_c_)_UB_, CI (KM_e_)_LB_, and CI (KM_e_)_UB_, HR represents HR, and CI represents CI (HR)_LB_ and CI (HR)_UB_. The red vertical line represents BF_10_ for the full Lung data.

**Figure A3.**
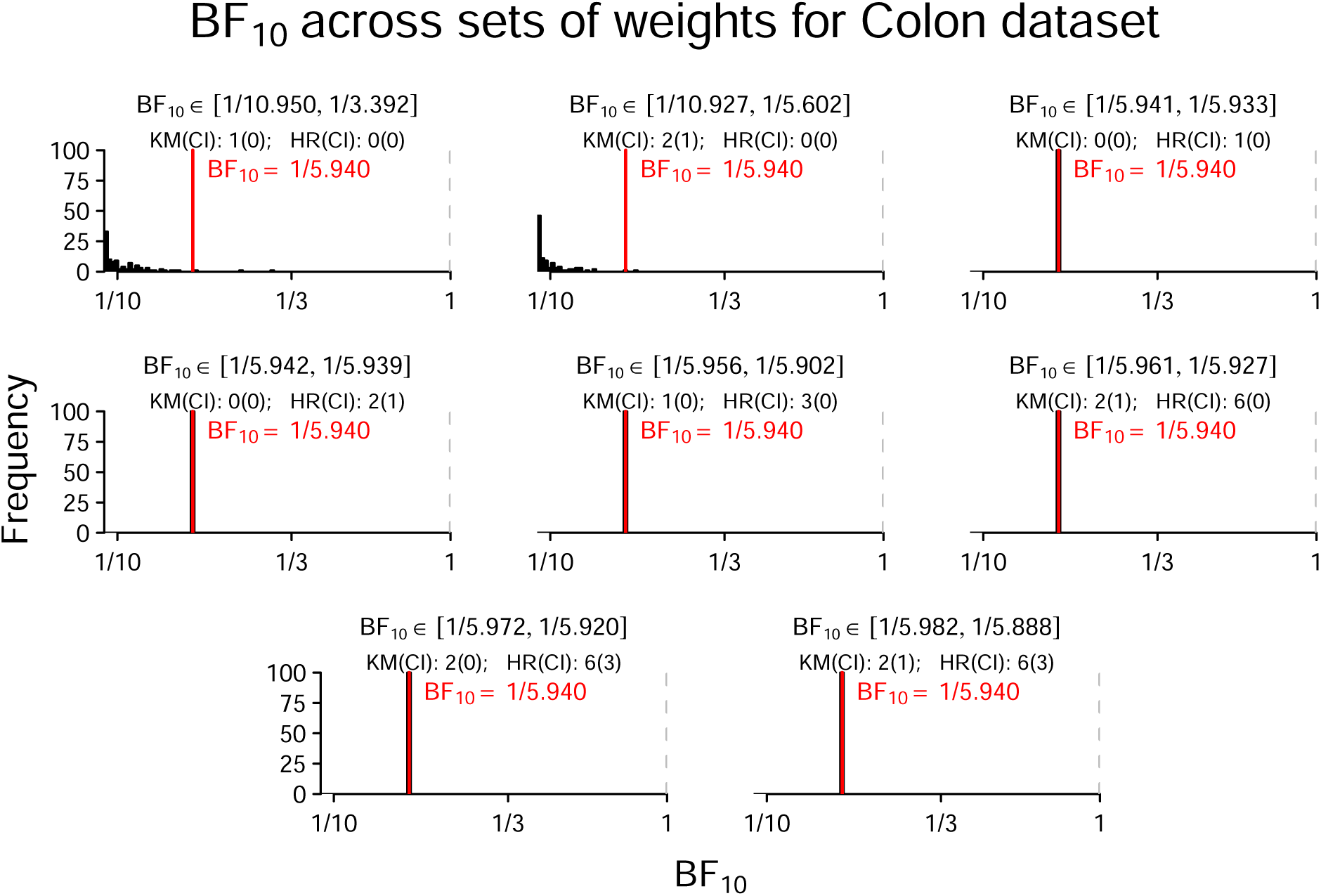
Distribution of BF_10_ for the Colon data set. Panels display BF_10_ for 100 simulated data sets using different sets of weights for summary statistics. The specific weights are printed in each panel, where KM represents KM_c_ and KM_e_, CI represents CI (KM_c_)_LB_, CI (KM_c_)_UB_, CI (KM_e_)_LB_, and CI (KM_e_)_UB_, HR represents HR, and CI represents CI (HR)_LB_ and CI (HR)_UB_. The red vertical line represents BF_10_ for the full Colon data.

## Appendix B

### Follow-up Analyses on the Influence of Different Weight Combinations

In the main manuscript, we have seen through simulations that using HR, CI (HR)*_LB_*, and CI (HR)*_U B_* as summary statistics yields the most accurate Bayes factors. We conducted follow-up simulations to investigate whether it is relevant which weights for HR, CI (HR)*_LB_*, and CI (HR)*_U B_* are chosen. Figure B1 shows the Bayes factors for 10 different weight settings for the Kidney, Lung, and Colon data sets. For each weight setting, 20 data sets were simulated with 200 PSO iterations and, accordingly, 20 Bayes factors were computed. It can be seen that it does not make a difference which specific weights are chosen, except in the situation where only HR is available, which diminishes the accuracy of the simulated Bayes factors.

**Figure B1.**
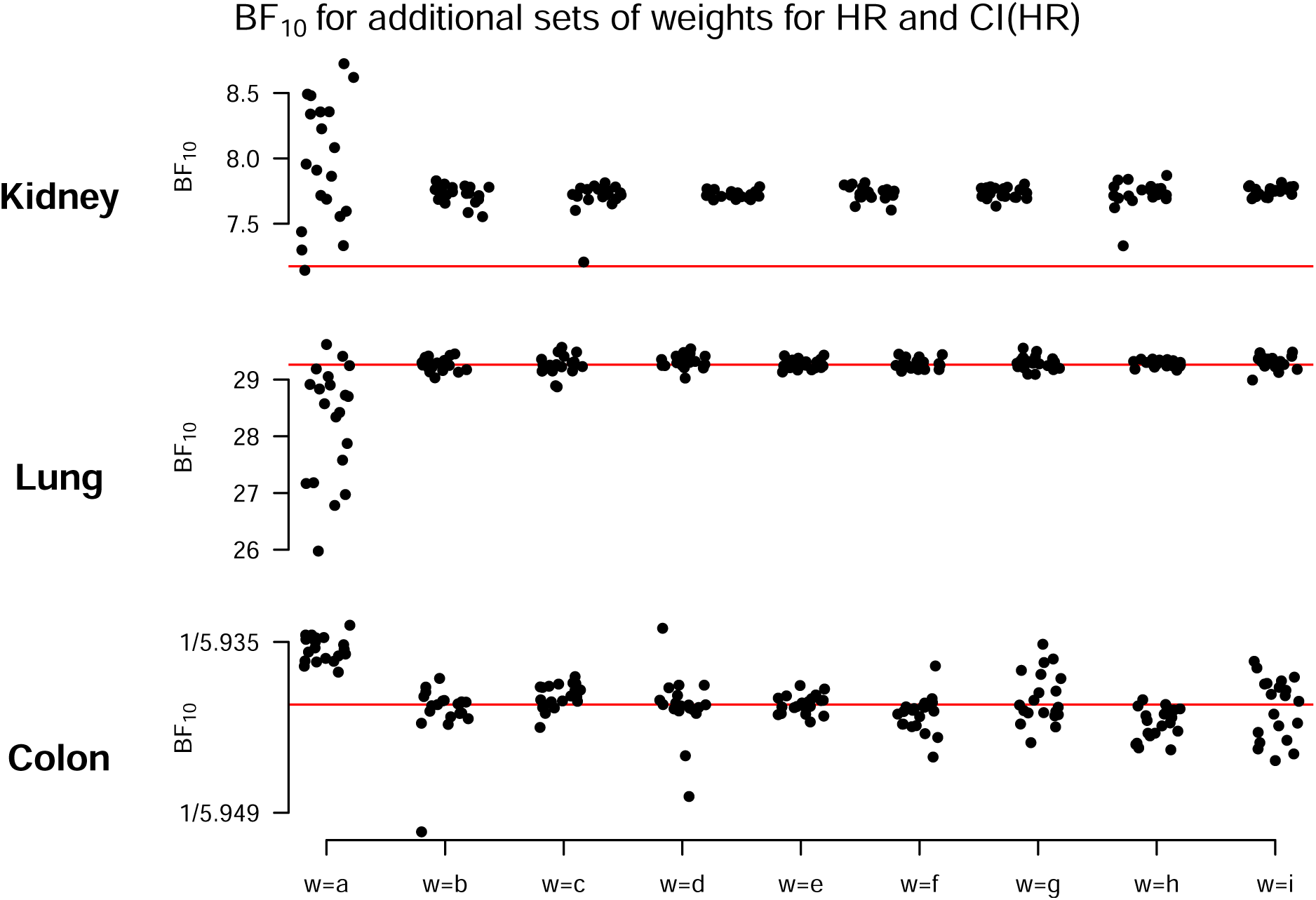
Accuracy of simulated Bayes factors as a function of weight combinations. The red horizontal line displays the true Bayes factor. The x-axis labels, matching the pattern HR [CI (HR)_LB_, CI (HR)_UB_ ], are: a=1[0, 0], b=2[1, 1], c=4[1, 1], d=1[2, 2], e=1[4, 4], f=3[1, 2], g=3[2, 1], h=1[2, 3], i=1[3, 2].

## Appendix C

### Results for Maximum Number of PSO Iterations

Figures C1, and C2 show the bias and variance of the simulated Bayes factors based on different maximum numbers of PSO iterations for the Kidney [54], Lung [53], and Colon [49] data sets from the “survival” R package [69], when using HR or a combination of HR and CI (HR), respectively. See main text for details.

**Figure C1.**
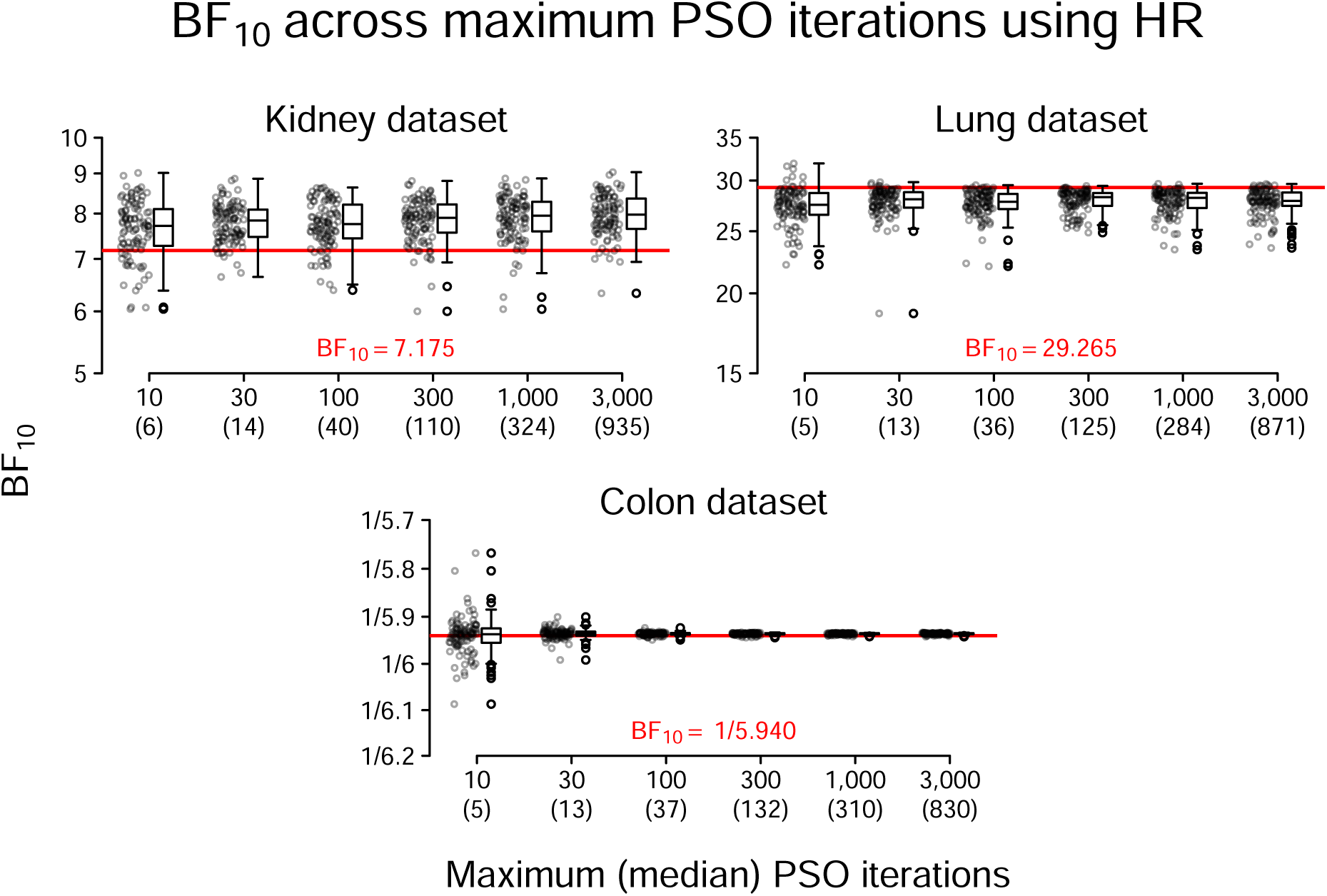
Distribution of BF_10_ applied to 100 simulated data sets using different maximum numbers of PSO iterations for the Kidney, Lung, and Colon data sets. HR is used for data simulation. Note that even though a maximum allowed number of PSO iterations was defined, optimization could stop earlier in case no improvement was found within one fifth of the maximum allowed number of iterations. Therefore, the median actual number of PSO iterations is given in parentheses on the x-axis. The red horizontal line represents BF_10_ for the actual data set.

**Figure C2.**
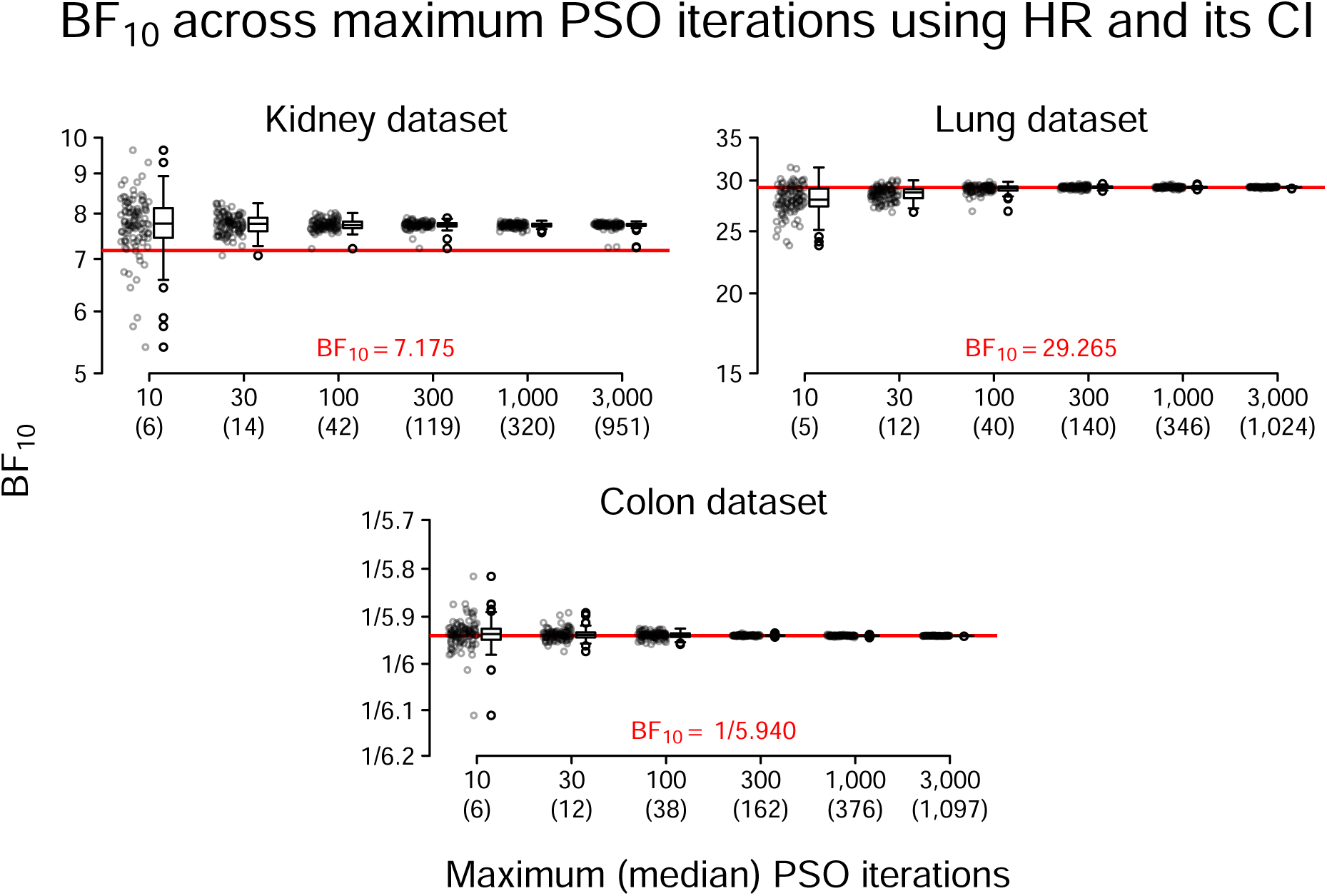
Distribution of BF_10_ applied to 100 simulated data sets using different maximum numbers of PSO iterations for the Kidney, Lung, and Colon data sets. HR and its 95%CI are used for data simulation. Note that even though a maximum allowed number of PSO iterations was defined, optimization could stop earlier in case no improvement was found within one fifth of the maximum allowed number of iterations. Therefore, the median actual number of PSO iterations is given in parentheses on the x-axis. The red horizontal line represents BF_10_ for the actual data set.

## Appendix D

### Run Time of Data Simulation and Bayes Factor Computation

Figure D1 shows the run time of our proposed procedure, where the process of data simulation is shown in the left panel and Bayes factor computation in the right panel. The run time was measured on a typical Windows 10 (*x*64) computer, with 16GB of RAM and an 11th Gen Intel(R) Core(TM) i7-1165G7 @ 2.80GHz CPU with 8 cores. For the measurement of the run times, we used the Kidney [54], Lung [53], and Colon [49] data sets that are available through the “survival” R package [69]. Furthermore, for data simulation, we either used 1, 2, or 4 CPU cores. For all data simulations, we simulated 20 data sets with exactly 100 PSO iterations. Both data simulation and computing Bayes factors were repeated 40 times to obtain a distribution of run times.

Figure D1 indicates that data simulation takes longer than computing Bayes factors. Using our settings, simulation of 20 data sets with 100 PSO iterations took approximately 60 − 150s, 70 − 180s, and 110 − 300s for the Kidney, Lung, and Colon data sets, respectively, using 1 − 4 CPU cores. Computing Bayes factors took approximately 4, 18, and 90s for the Kidney, Lung, and Colon data sets, respectively. It can be seen that both data simulation and Bayes factor computation take longer the larger the sample size of the underlying data set is. Moreover, it is visible that the run time for data simulation can be drastically reduced by using multiple CPU cores. However, this is only possible when more than one data set is to be simulated. Ideally, the number of data sets to be simulated is a multiple of the number of used CPU cores. Further, parallel processing with multiple CPU cores is only beneficial when a considerable number of PSO iterations is used and when the sample size of the underlying data set is not too small. Otherwise, the initialization of the CPU cores takes up too much time for the benefits of parallel processing to unfold.

**Figure D1.**
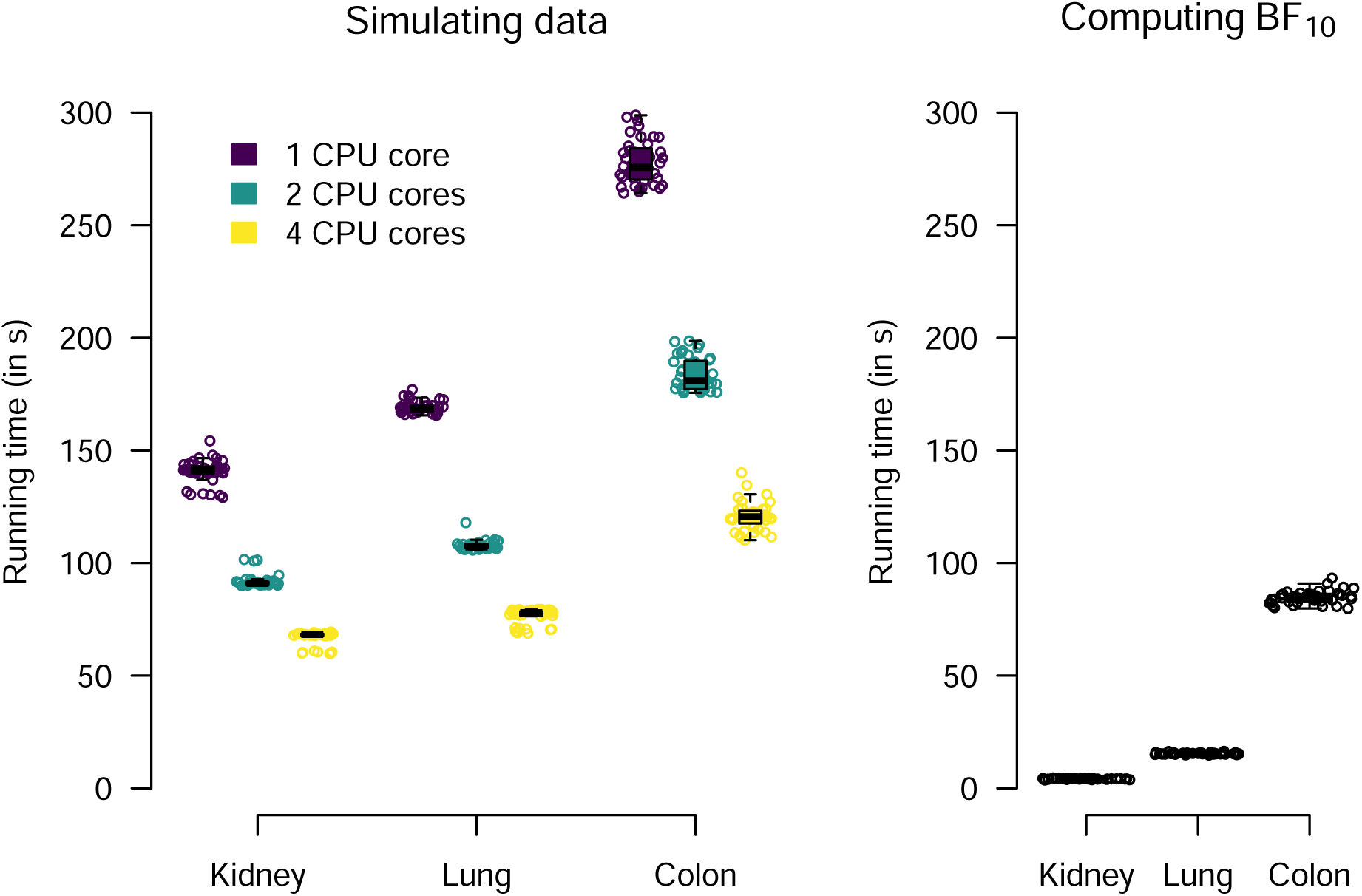
Runtime for data simulation of 20 data sets (left panel) and for Bayes factor computation (right panel) using the Kidney, Lung, and Colon data sets. For each data set, data simulation and Bayes factor computation was repeated 40 times. For data simulation (left panel), run times with 1, 2, and 4 CPU cores are shown in purple, green, and yellow, respectively.

## Appendix E

### Exploration of Factors Contributing to the Accuracy Difference Between Our Method and the Method by [4]

In the main manuscript we found that our method for simulating Bayes factors provides more accurate Bayes factors than the method by [4] (see Figure 2). To explore possible reasons for this discrepancy, we conducted further simulations.

We took the Kidney, Lung, and Colon data sets and generated data sets of variable sizes through bootstrapping (i.e., sampling with replacement). The sizes of the bootstrapped data sets were *n* = {40, 80, 160, 320, 640, 1280}. This process was repeated 10 times. Thus, in total we have 3 × 6 × 10 = 180 bootstrapped data sets. Subsequently, two separate analyses were conducted.

First, for each bootstrapped data set, we used the full data to compute a Bayes factor using our method. This Bayes factor served as the “truth”. Moreover, we applied our approach and the approach by [4] on each bootstrapped data set. In other words, we have (1) the “true” Bayes factor, (2) the median Bayes factor resulting from our approach (median because we simulate 8 data sets), and (3) the Bayes factor resulting from the method by [4] for each bootstrapped data set. Then, for each bootstrapped data set we computed two numbers: 1) *log* (*BF*_Our_*/BF*_True_) and 2) *log* (*BF*_Bartos_*/BF*_True_). Figure E1 shows these accuracy ratios as a function of the sample size of the bootstrapped data sets. For the Kidney and Lung data sets, the accuracy of the approach by [4] decreases as the sample size increases. For the Colon data set, accuracy is almost equal across sample sizes for the approach by [4], except for small sample sizes, where less accuracy is observed. The accuracy of the Bayes factors resulting from the summary BF procedure are good for all data sets and for almost all sample sizes. Only for the Kidney data set it was observed that accuracy diminishes for higher sample sizes.

Second, we explored whether the shape of the likelihood influences the accuracy of the resulting Bayes factors. For our method, we computed the normalized Efron approximation to the true log Cox partial likelihood and for the method by [4] we computed the normalized log Normal likelihood (i.e., a log Normal density) on a fine grid of *β*values for each bootstrapped data set. The normalization was done to make the log likelihoods of the two methods comparable. Subsequently, we computed the similarity of the two normalized log likelihood distributions by using the Kullback-Leibler divergence (KLD; [6]). The results are shown in Figure E2. It can be seen that in most cases, the KLD is very low, indicating that the distributions were very similar. As the sample size increased, the KLD decreased, indicating higher similarity of the normalized log likelihoods for larger samples.

**Figure E1.**
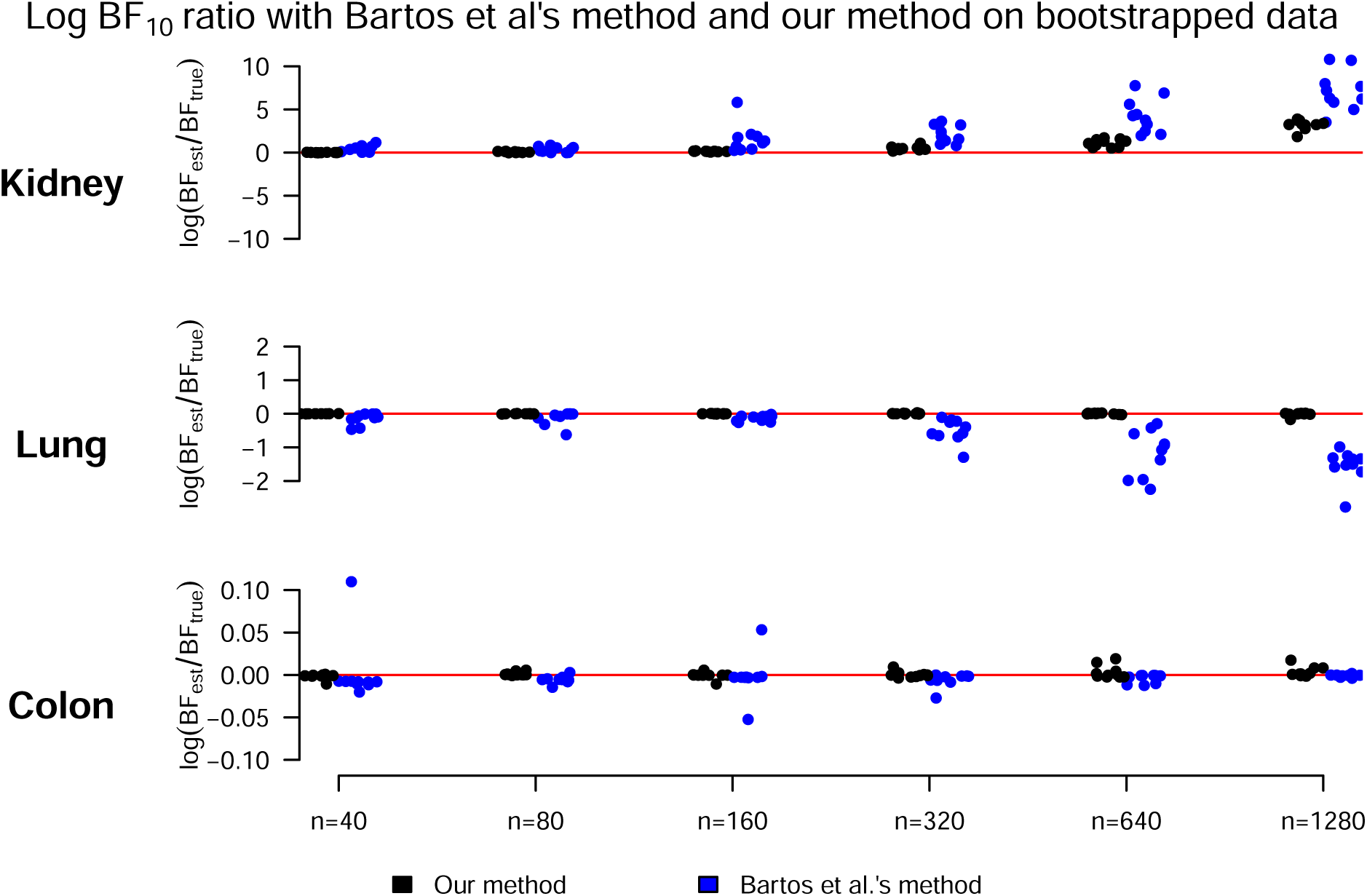
Log ratio of the estimated Bayes factor using our method (black) or the method by [4] (blue) and the true Bayes factor as a function of the sample size of the bootstrapped data sets. The red horizontal line shows the optimum.

**Figure E2.**
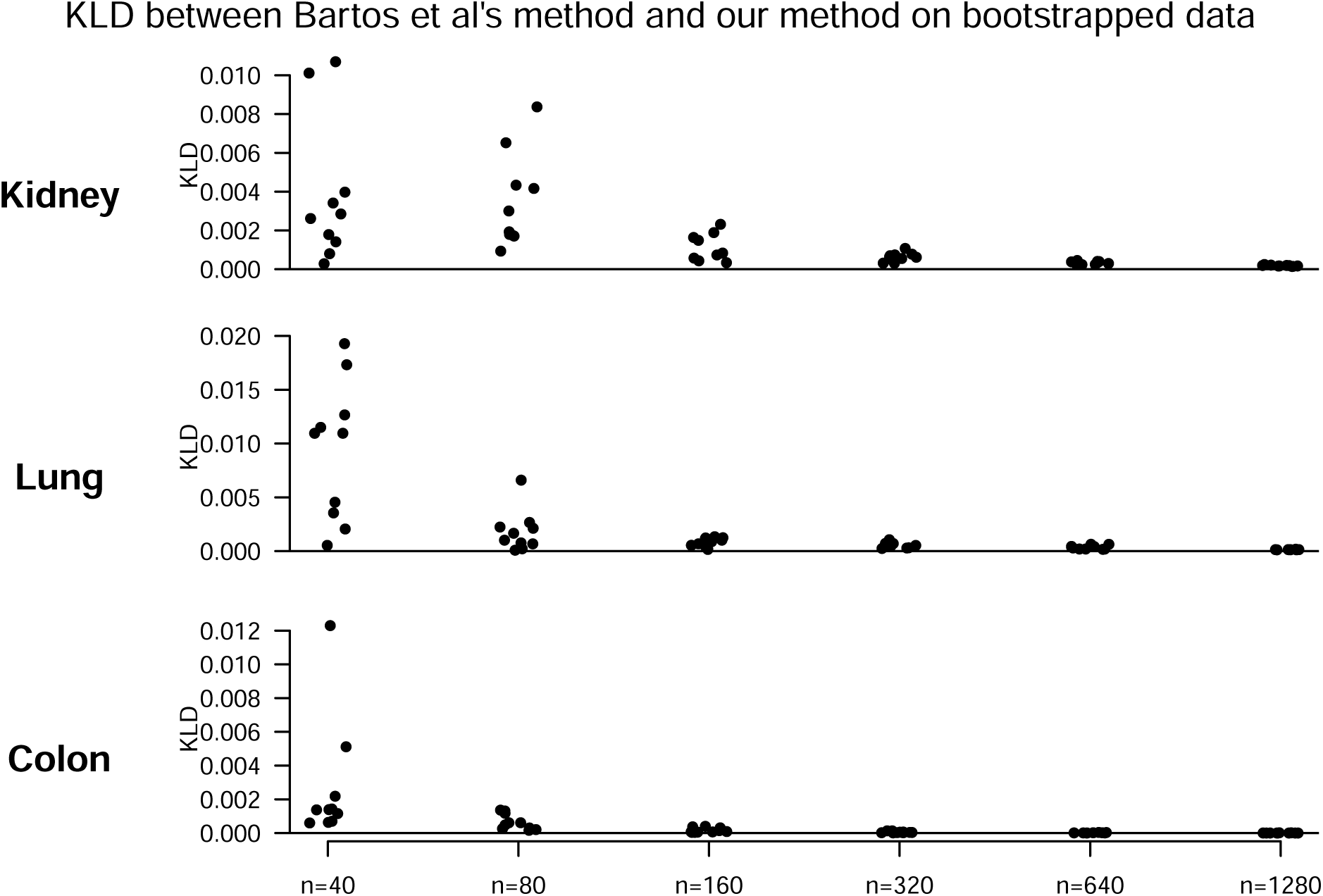
Kullback-Leibler divergence (KLD) between the normalized likelihood of our method and the normalized likelihood of the method by [4] as a function of the sample size of bootstrapped data sets.

